# Quantifying the impact of early-life growth adversity on later-life health

**DOI:** 10.1101/2025.06.16.25327714

**Authors:** Raphael Goldman-Pham, Matthew P. Alter, Rebecca Bao, Sophie É. Collins, Catherine L. Debban, James P. Allinson, Antony Ambler, Alain G. Bertoni, Avshalom Caspi, Stephanie Lovinsky-Desir, Magnus P. Ekstrom, James C. Engert, David R. Jacobs, Daniel Malinsky, Ani Manichaikul, Erin D. Michos, Terrie E. Moffitt, Elizabeth C. Oelsner, Sandhya Ramrakha, Stephen S. Rich, Coralynn Sack, Sanja Stanojevic, Padmaja Subbarao, Karen Sugden, Reremoana Theodore, Karol E. Watson, Benjamin Williams, Bin Yang, Josée Dupuis, Seif O. Shaheen, R. Graham Barr, Robert J. Hancox, Benjamin M Smith

**Author notes:** Corresponding author: Benjamin M Smith Research Institute of the McGill University Health Centre 5252 boulevard de Maisonneuve West Montreal, Quebec H4A 3S9 Canada Columbia University Medical Center 630 W 168^th^ Street, PH9-105 New York, NY 100032 United States of America.

## Abstract

**Background:** Early-life growth adversity is important to later-life health, but precision assessment in adulthood is challenging. We evaluated whether the difference between attained and genotype-predicted adult height (“height-GaP”) would associate with prospectively ascertained early-life growth adversity and later-life all-cause and cardiovascular mortality.

**Methods:** Data were first analyzed from the Avon Longitudinal Study of Parents and Children (ALSPAC) and UKBiobank. Genotype-predicted height was calculated using a multi-ancestry polygenic height score. Height-GaP was calculated as the difference between measured and genotype-predicted adult height. Early-life growth conditions were ascertained prospectively via standardized procedures (ALSPAC) and mortality via death register (UKBiobank). Regression models adjusted for age, sex, genotype-predicted height and genetic ancestry. Analyses were replicated in the Dunedin Multidisciplinary Health and Development Study (DMHDS) and the Multi-Ethnic Study of Atherosclerosis (MESA).

**Findings:** Among 4,582 ALSPAC participants (median [IQR] age: 24[18-25] years at height-GaP assessment), lower gestational age at birth, greater pre- and post-natal deprivation indices, tobacco smoke exposure and less breastfeeding were associated with larger adult height-GaP deficit (p<0.01). Among 483,385 UKBiobank participants (mean±SD age: 56±8 years at height-GaP assessment), height-GaP deficit was associated with death from all-causes (adjusted hazard ratio comparing highest-to-lowest height-GaP deficit quartile [aHR]:1.25 95%CI:1.21-1.29), atherosclerotic cardiovascular disease (aHR:1.33 95%CI:1.24-1.43) and coronary heart disease (aHR:1.68 95%CI:1.52-1.86). Early- and later-life height-GaP associations replicated in DMHDS and MESA.

**Interpretation:** This study introduces a simple index of early-life growth adversity deployable in adulthood to investigate the developmental origins of longevity and improve health equity across the life course.

**Funding:** Federal agencies and academic institutions.

## INTRODUCTION

The period of human ontogenetic growth, which spans conception to late adolescence, is increasingly recognized as an important window of susceptibility to exposures and events that influence later-life health.^1^ Transforming this understanding into substantive reductions in disease burden and health inequity is challenging, in part, due to the lack of simple methods to quantify early-life growth adversity in adulthood. These challenges include the diverse and often correlated nature of early-life factors affecting growth, recall and selection bias when ascertained retrospectively, and the long latency to later-life health outcomes. This study sought to evaluate a novel quantitative index of early-life growth adversity deployable in adulthood to facilitate life course research on the developmental origins of health and disease.

Human height increases during the period of ontogenetic growth and is determined in part by genetics and by early-life growth conditions. A recent genome-wide association study of ∼5.4 million adults reported a saturated map of common genetic variants associated with adult height that accounts for over 90% of trait heritability and up to 45% of trait variance.^2^ It follows that the difference between measured and genotype-predicted adult height, here referred to as “height-GaP”, may represent a simple index of early-life growth adversity. If true, such an index could i) facilitate discovery of threats to human development, ii) serve as a surrogate endpoint for early-life interventions aiming to improve later-life health, and iii) improve adult risk stratification and endo-phenotyping for diseases with developmental origins.

This study’s first objective was to determine whether prospectively ascertained early-life factors known to influence ontogenetic growth are associated with adult height-GaP. The second objective was to determine whether adult height-GaP is associated with later-life mortality from all-causes and from atherosclerotic cardiovascular disease, the latter being a major public health burden with established developmental origins.^1,3–7^

## METHODS

### Design

Cross-sectional and longitudinal analyses of cohort data.

### Data

The Avon Longitudinal Study of Parents and Children (ALSPAC) is a prospective birth cohort that enrolled women who were pregnant in Avon, England with expected delivery dates between April 1 1991 and December 31 1992.^8–10^ Initial recruitment of 14,541 pregnancies resulted in 13,988 children alive after 1 year. Following further recruitment of eligible participants, 15,447 pregnancies were included in the study, with 14,901 children alive after 1 year. Parents and children have been characterized over the past four decades including anthropometry, questionnaires and genotyping. ALPAC provides a searchable data dictionary online.^11^ For this analysis, we included participants with measured standing height performed at visit year 18 or 24, genotype data and information on at least one prospectively ascertained early-life growth condition (n=4,582). Ethical approval for the study was obtained from the ALSPAC Ethics and Law Committee and the Local Research Ethics Committees.

UKBiobank (UKB) is a prospective cohort that enrolled over 500,000 adults aged 40-69 years between 2006 to 2010 across England, Scotland and Wales.^12^ Participants were extensively characterized at baseline including standardized anthropometry, questionnaires, genotyping, with follow-up assessment of vital status. For this analysis, we included participants with measured adult standing height, genotype data and information on vital status (n=483,385).

The Dunedin Multidisciplinary Health and Development Study (DMHDS) is a population-based birth cohort of participants born between April 1972 and March 1973 at Queen Mary Maternity Hospital in Dunedin, New Zealand.^13,14^ Participants were enrolled at 3 years old and have undergone periodic standardized assessments into the fifth decade of life. For this analysis, we included participants with measured adult standing height, genotype data, and at least one prospectively ascertained early-life growth condition (n=855).

The Multi-Ethnic Study of Atherosclerosis (MESA) is a United States multi-site cohort that enrolled 6,814 adults 45 to 84 years old between 2000 and 2002 who self-identified as White, African American, Hispanic or Chinese. Exclusion criteria included clinically apparent cardiovascular disease and impediments to long-term follow-up. For this analysis, we included participants with measured standing height at baseline, genotype data, and information on vital status (n=6,352).

Institutional research ethics board approval was obtained and participants provided written informed consent in all studies.

### Measured standing height

All cohorts measured standing shoeless height according to standardized protocols with calibrated stadiometers.

### Genotyping

ALSPAC genotyping was performed via Illumina 660 quad sequencing with imputation according to the 1000 Genomes reference panel.^9^ UKBiobank genotyping was performed using the UKBiobank Axiom Array with imputation according to a combination of the Haplotype Reference Consortium (HRC) and 1000 Genomes reference panels.^12^ DMHDS genotyping was performed using the Illumina HumanOmni Express 12 BeadChip array with imputation using the 1000 Genomes phase 3 reference panel.^15^ MESA genotyping was obtained through TOPMed Freeze 10b, with quality control performed based on existing TOPMed protocols.^16^

### Genotype-predicted adult height

Genotype-predicted height was computed using the all-ancestry polygenic height weights reported by Yengo et al.^2^ Each participant’s polygenic height score was first calculated as the weighted sum of height-increasing alleles (additive model; missing effect allele status assigned the sample effect allele prevalence). Genotype-predicted height was then computed by fitting cohort- and sex-specific linear regression models of measured height. Ancestry-specific polygenic height scores^2^ were also computed using self-reported race-ethnic group for sensitivity analyses.

### Height-GaP

Height-GaP was computed as the difference of measured adult height minus genotype-predicted height in centimeters. A lower height-GaP value represents a larger deficit in measured height compared to genotype-predicted height.

### Early-life growth conditions in ALSPAC and DMHDS

Birthweight was extracted from medical records in ALSPAC and DMHDS; birth length was measured by trained ALSPAC staff and extracted from medical records in DMHDS.^17,18^ Gestational age was estimated from the date of last menstrual period and confirmed via ultrasound in ALSPAC, and extracted from medical records in DMHDS.^17^ Standardized questionnaire items were used to assess breastfeeding status and duration (months), maternal smoking during pregnancy (cigarettes/day) and household tobacco smoke exposure in ALSPAC and DMHDS.^8,19^ Dietary patterns in ALSPAC were assessed using standardized questionnaires and summarized using principal components analysis.^20^ Residential address information was used to compute neighbourhood-level indices of multiple deprivation during pregnancy, birth and the average across post-natal ages 1-12 years in ALSPAC, with higher values corresponding to greater deprivation.^21^ In DMHDS, socioeconomic status was quantified with a six-point occupational measure that was most widely used in the New Zealand research community while the participants were growing up.^22^ The variable used in our analyses, childhood socioeconomic status, is the average of the highest socioeconomic status level of either parent, assessed repeatedly at the participant’s birth and at ages 3, 5, 7, 9, 11, 13, and 15 years.

### Mortality in UKBiobank and MESA

Information on vital status was ascertained in UKBiobank via linkage to the British National Death Registry and in MESA via 9-12 month interval participant residence telephone contacts and linkage to the National Death Index of the National Vital Statistics System yielding date of death and the International Classification of Disease- (ICD)-10 code listed as the primary (underlying) cause of death.^23,24^ Time until death or censorship was computed as the difference in years between death or last study contact and baseline study visit.

Deaths due to atherosclerotic cardiovascular disease (fatal coronary heart disease or fatal stroke) and atherosclerotic coronary heart disease were defined in UKBiobank based on the ICD-10 code listed as the underlying (primary) cause of death (I20-25, I60-61, I63-64)^25^ and in MESA by standardized adjudication that included paired cardiologist or neurologist review of abstracted medical records, with disagreements resolved by full committee review.^26^

### Other variables (See Supplement Table 1 for additional details)

Age, sex and race or ethnicity were self-reported. Principal components of genetic ancestry were derived from genotype data in UKBiobank, DMHDS and MESA.^12^ The ALSPAC sample consisted only of persons of European ancestry defined by genetic principal components.^27^ In UKBiobank and MESA, cigarette smoking status, pack-years of smoking, alcohol use status, average number of drinks per week, quantity of moderate-to-vigorous physical activity per week, household income, educational attainment and health insurance status were derived from standardized questionnaire items. Diabetes status was defined by a fasting blood glucose ≥7.0 mmol/L (126 mg/dl) or diabetes medication use, low-density lipoprotein (LDL) cholesterol concentration was derived from fasting blood sample triglyceride concentration. Hypertension status was defined by standardized automated measures of systolic and diastolic blood pressure or the use of anti-hypertensive medication.

For height loss sensitivity analyses, annualized longitudinal change in measured height was computed in UKBiobank as the difference in height in cm between baseline and follow-up assessment divided by the time interval between assessments in years.

### Statistical analysis

Participant characteristics are summarized by quantile of adult height-GaP. The hypothesized causal relationships of early-life growth conditions with later-life health outcomes are depicted in a directed acyclic graph (*Supplemental Figure 1).* Associations were first computed using ALSPAC and UKBiobank data, followed by replication analyses in DMHDS and MESA.

Separate generalized linear and spline regression models of adult height-GaP were fit for each prospectively ascertained early-life growth condition. Models were unadjusted and adjusted for age at height-GaP assessment, sex and genotype-predicted height. Tabular results are reported per 1-SD or quantile contrast depending on whether the linear or spline model fit was superior. The statistical significance threshold for ALSPAC analyses was a two-sided p-value=0.010 to account for testing of five early-life growth categories (nutrition [breastfeeding, diet], peri-natal [gestational age at birth, birth weight, birth length], socio-economic deprivation [multiple deprivation index], and noxious exposures [tobacco smoke, PM_2.5_). Missing data were imputed using polytomous logistic regression, and predictive mean matching, for categorical and continuous variables respectively; the threshold in the DMHDS replication analysis was a two-sided p-value=0.050.

Proportional hazard models of mortality (all-cause and cause-specific) were fit with baseline height-GaP as the independent variable of interest. Model 1 adjusted for age, sex and principal components of genetic ancestry. Model 2 (main model) additionally adjusted for genotype-predicted height to provide the model with a linear combination of information equivalent to measured height. Heterogeneity of associations by sex and by race-ethnicity were evaluated by including product terms in regression models. The statistical significance threshold in UKBiobank was a two-sided p-value=0.017 (0.05/3) to account for testing three mortality outcomes; the threshold in the MESA replication analysis was a two-sided p-value=0.050. Mortality associations with measured height and genotype-predicted height were also assessed using the aforementioned models.

Sensitivity analyses: To evaluate later-life height loss we corrected each participant’s measured height (and thus height-GaP) back to the age of height loss onset using i) the median annualized height loss and the participant’s age and sex, and ii) a ‘worst case’ scenario of height-loss confounding (See *Supplemental Methods* for details).

To account for potential confounding of the height-GaP association with later-life mortality by adult health-related factors, the main model was additionally adjusted for the following: baseline hypertension status, systolic blood pressure, diabetes status, weight status, cigarette smoking status (current, former, never), pack-years of smoking, alcohol consumption status and drink frequency, minutes of weekly moderate and weekly vigorous physical activity, educational attainment, household income and, in MESA, health insurance status. We note, however, that these adult health conditions have been implicated in the developmental origin of health and disease paradigm and may represent partial or full mediators.^1^

Finally, ALSPAC analyses were repeated among participants with complete data and UKBiobank and MESA analyses repeated using ancestry-specific polygenic height scores.

All analyses were performed using R version 4.4.1 and Python version 3.12.5.

## RESULTS

Characteristics of the 4,582 ALSPAC participants included in this analysis are summarized in **Table 1**. The median (IQR) age at the time of height-GaP assessment was 24 (18, 25) years, 56.3% were female and the mean±SD measured height was 180±7 cm for males and 166±6 cm for females. The polygenic height score accounted for 40.3% and 37.6% of the variance in adult height among males and females respectively, and the mean±SD height-GaP was 0.0±5.0 cm (95^th^ percentile range: -9.9 to 9.7 cm). Characteristics of excluded participants are summarized in *Supplemental Table 2*.

**Table 1:** Characteristics of participants included in the ALSPAC analyses.

|  | **All** | **By Height-GaP Quartile** |  |  |  |
| --- | --- | --- | --- | --- | --- |
|  |  | **1** | **2** | **3** | **4** |
| No. | 4,582 | 1,146 | 1,146 | 1,145 | 1,145 |
| Sex, no. (%) |  |  |  |  |  |
| Female | 2,579 (56.3) | 643 (56.1) | 656 (57.2) | 649 (56.7) | 631 (55.1) |
| Male | 2,003 (43.7) | 503 (43.9) | 490 (42.8) | 496 (43.3) | 514 (44.9) |
| Age at height-GaP assessment, years (IQR) | 24 (18, 25) | 24 (18, 25) | 24 (18, 25) | 24 (19, 25) | 24 (18, 25) |
| Genotype-predicted height cm | 171.9 (7.9) | 172.0 (7.9) | 171.6 (7.9) | 171.7 (8.0) | 172.1 (7.9) |
| Height-GaP, cm | 0.0 (5.1) | -6.4 (2.7) | -1.6 (0.9) | 1.6 (0.9) | 6.4 (2.6) |
| Height, cm | 171.9 (9.4) | 165.6 (8.1) | 170.0 (7.8) | 173.3 (8.0) | 178.5 (8.6) |
| Pregnancy index of multiple deprivation quintile, mean | 2.91 (1.38) | 3.04 (1.40) | 2.94 (1.36) | 2.91 (1.37) | 2.78 (1.36) |
| Maternal smoking during pregnancy, no. (%) |  |  |  |  |  |
| Never | 3,751 (87.3) | 988 (83.6) | 931 (87.1) | 944 (87.9) | 977 (90.5) |
| 1-4 cigarettes/day, n (%) | 144 (3.4) | 46 (4.3) | 41 (3.8) | 31 (2.9) | 26 (2.4) |
| 5-9 cigarettes/day, n (%) | 126 (2.9) | 39 (3.6) | 33 (3.1) | 34 (3.2) | 20 (1.9) |
| 10-19 cigarettes/day, n (%) | 209 (4.9) | 63 (5.9) | 51 (4.8) | 51 (4.7) | 44 (4.1) |
| 20+ cigarettes/day, n (%) | 67 (1.6) | 28 (2.6) | 13 (1.2) | 14 (1.3) | 12 (1.1) |
| “Healthy” diet principal component during pregnancy | 0.24 (0.95) | 0.14 (0.96) | 0.24 (0.95) | 0.26 (0.95) | 0.34 (0.92) |
| Gestational age at birth, weeks | 39.5 (1.8) | 39.3 (2.0) | 39.5 (1.8) | 39.5 (1.8) | 39.5 (1.7) |
| Birth length, cm | 50.6 (2.1) | 49.9 (2.1) | 50.4 (2.1) | 50.8 (2.0) | 51.4 (2.0) |
| Birth weight, kg | 3.43 (0.53) | 3.26 (0.57) | 3.40 (0.52) | 3.47 (0.50) | 3.57 (0.50) |
| Breastfeeding duration, no. (%) |  |  |  |  |  |
| Never | 641 (15.8) | 192 (19.1) | 166 (16.5) | 158 (15.6) | 125 (12.1) |
| <1 month | 580 (14.3) | 158 (15.7) | 136 (13.5) | 143 (14.1) | 143 (13.9) |
| 1-<3 months | 623 (15.4) | 159 (15.8) | 157 (15.6) | 159 (15.7) | 148 (14.4) |
| 3-<6 months | 601 (14.8) | 138 (13.7) | 145 (14.4) | 151 (14.9) | 167 (16.2) |
| 6+ months | 1608 (39.7) | 358 (35.6) | 401 (39.9) | 401 (39.6) | 448 (43.5) |
| Childhood index of multiple deprivation quintile, mean | 2.77 (1.26) | 2.89 (1.29) | 2.79 (1.24) | 2.77 (1.27) | 2.64 (1.24) |
| Childhood household tobacco smoke exposure duration, no. (%) |  |  |  |  |  |
| 0 hours/week | 1,813 (54.2) | 388 (48.2) | 446 (54.6) | 476 (55.4) | 503 (58.2) |
| >0-5 hours/week | 984 (29.4) | 241 (29.9) | 238 (29.1) | 254 (29.6) | 251 (29.0) |
| >5-10 hours/week | 177 (5.3) | 56 (7.0) | 47 (5.8) | 41 (4.8) | 33 (3.8) |
| >10-20 hours/week | 189 (5.6) | 61 (7.6) | 45 (5.5) | 45 (5.2) | 38 (4.4) |
| 20+ hours/week | 183 (5.5) | 59 (7.3) | 41 (5.0) | 43 (5.0) | 40 (4.6) |
| “Healthy” diet principal component at 38 months | 0.05 (0.99) | -0.01 (0.91) | 0.07 (0.99) | 0.06 (1.00) | 0.08 (1.03) |
| Residential outdoor PM<sub>2.5</sub> concentration, µg/m<sup>3</sup> | 13.26 (0.77) | 13.24 (0.74) | 13.28 (0.75) | 13.27 (0.79) | 13.25 (0.79) |

Mean (SD) unless otherwise specified.
Abbreviations: ALSPAC = Avon Longitudinal Study of Parents and Children; PM<sub>2.5</sub> = particulate matter <2.5 micrometres in diameter.

Characteristics of the 483,385 UKBiobank participants included are summarized in **Table 2**. The mean±SD age at height-GaP assessment was 56±8 years, 54% were female, mean±SD measured height was 176±7 cm for males and 163±4 cm for females, 10.5% were current smokers and 34.5% were former smokers (median [IQR] 19 [10, 32] pack-years among ever smokers). The polygenic height score accounted for 37.1% and 35.2% of variance in measured height among males and females respectively and the mean±SD height-GaP was 0.0±5.2 cm (95^th^ percentile range: -10.1 to 10.5 cm). Over a median of 12.4 years of follow-up (5,996,608 person-years) there were 35,703 deaths, of which 7,177 were attributed to atherosclerotic cardiovascular disease and 3,801 to atherosclerotic coronary heart disease. Participant characteristics were generally similar across height-GaP deficit quartile, except those with larger height-GaP deficit tended to be older, to have ever smoked and to have a higher number of pack-years. Excluded participant characteristics are summarized in *Supplemental Table 3*.

**Table 2:**
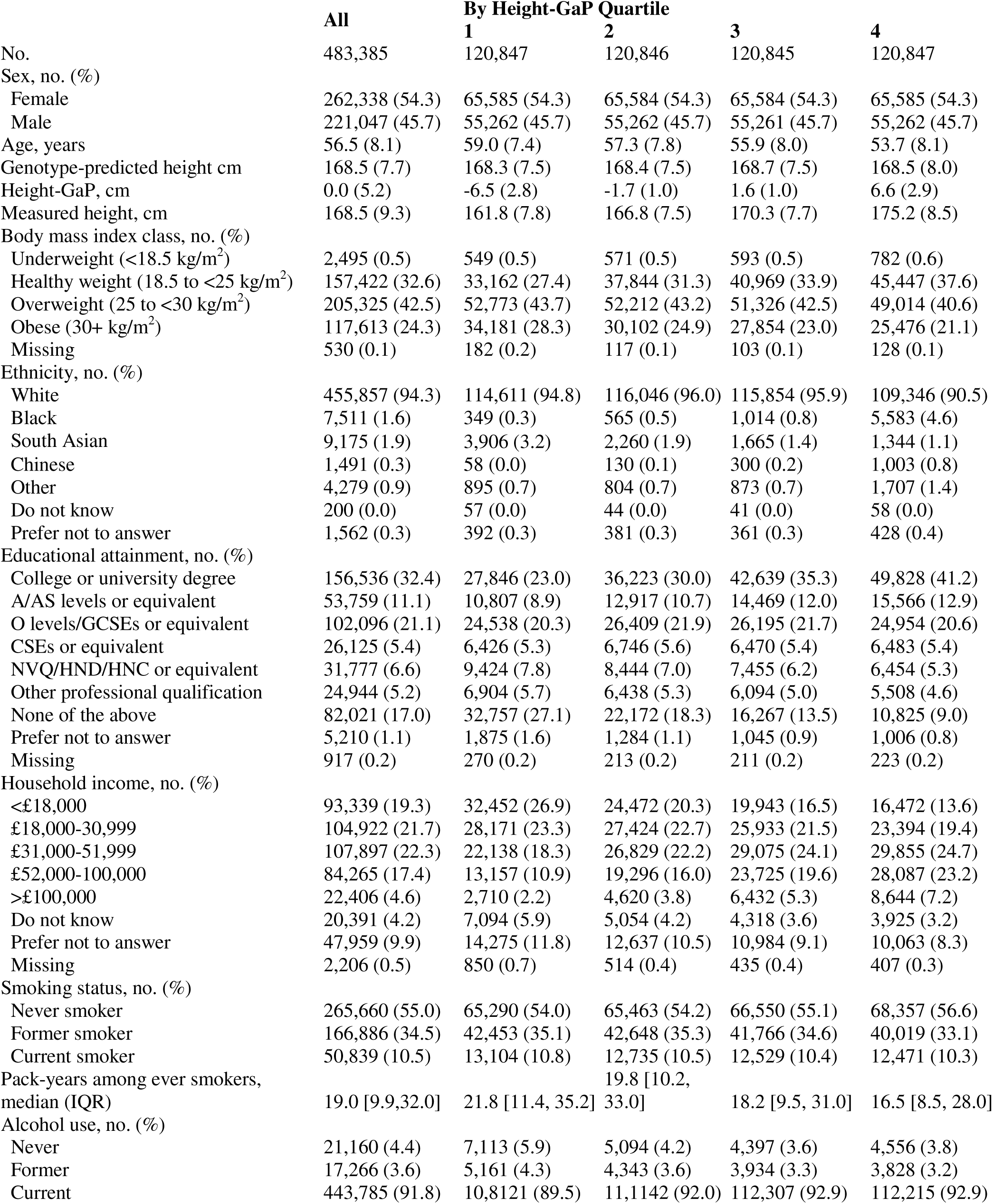
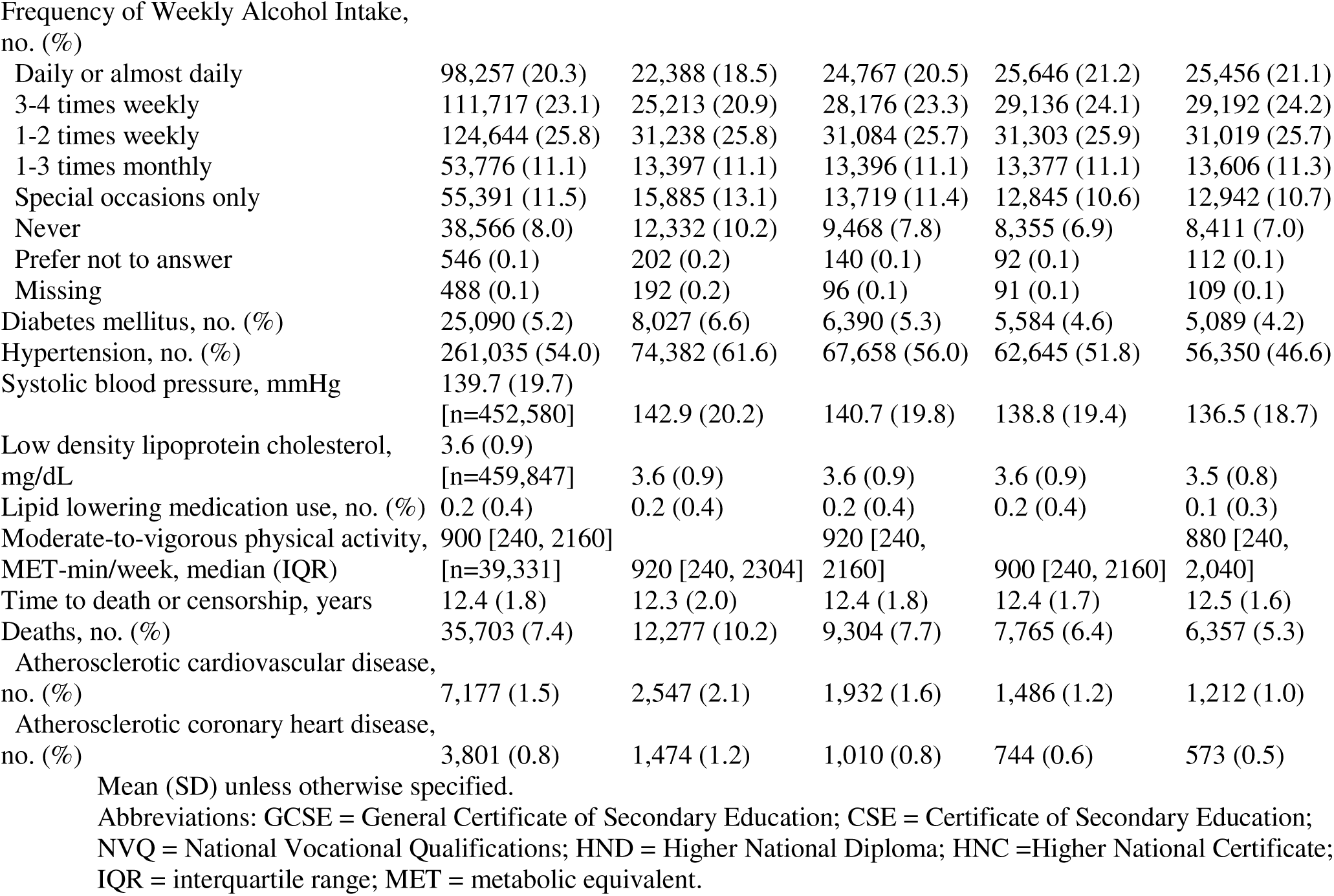
Characteristics of participants included in the UKBiobank analyses.

Characteristics of the replication cohorts are presented in *Supplemental Tables 4*-*5*. The DMHDS sample included 855 participants (age: 26±1 years at height-GaP assessment; 49% female; height: 178±6 cm for males and 165±6 cm for females) with a height-GaP of 0.0±5.3 cm (95^th^ percentile range: -9.9 to 10.3 cm). The MESA sample included 6,352 participants (age: 62±10 years at height-GaP assessment; 52.3% female; measured height: 173±8 cm for males and 160±7 cm for females) with a height-GaP of 0.0±5.7 cm (95^th^ percentile range: -11.1 to 11.5 cm). Self-identified race-ethnic proportions were 39.1% White, 26.2% African American, 22.6% Hispanic, and 12.1% Chinese. Over a median of 15.0 years of follow-up (92,319 person-years) there were 1,337 deaths, of which 233 and 155 were adjudicated as being due to atherosclerotic cardiovascular and coronary heart disease, respectively.

### Early-life growth conditions and adult height-GaP

The adjusted associations of early-life growth conditions with adult height-GaP in ALSPAC are summarized in **Figure 1** and *Supplemental Table 6*.

**Figure 1:**
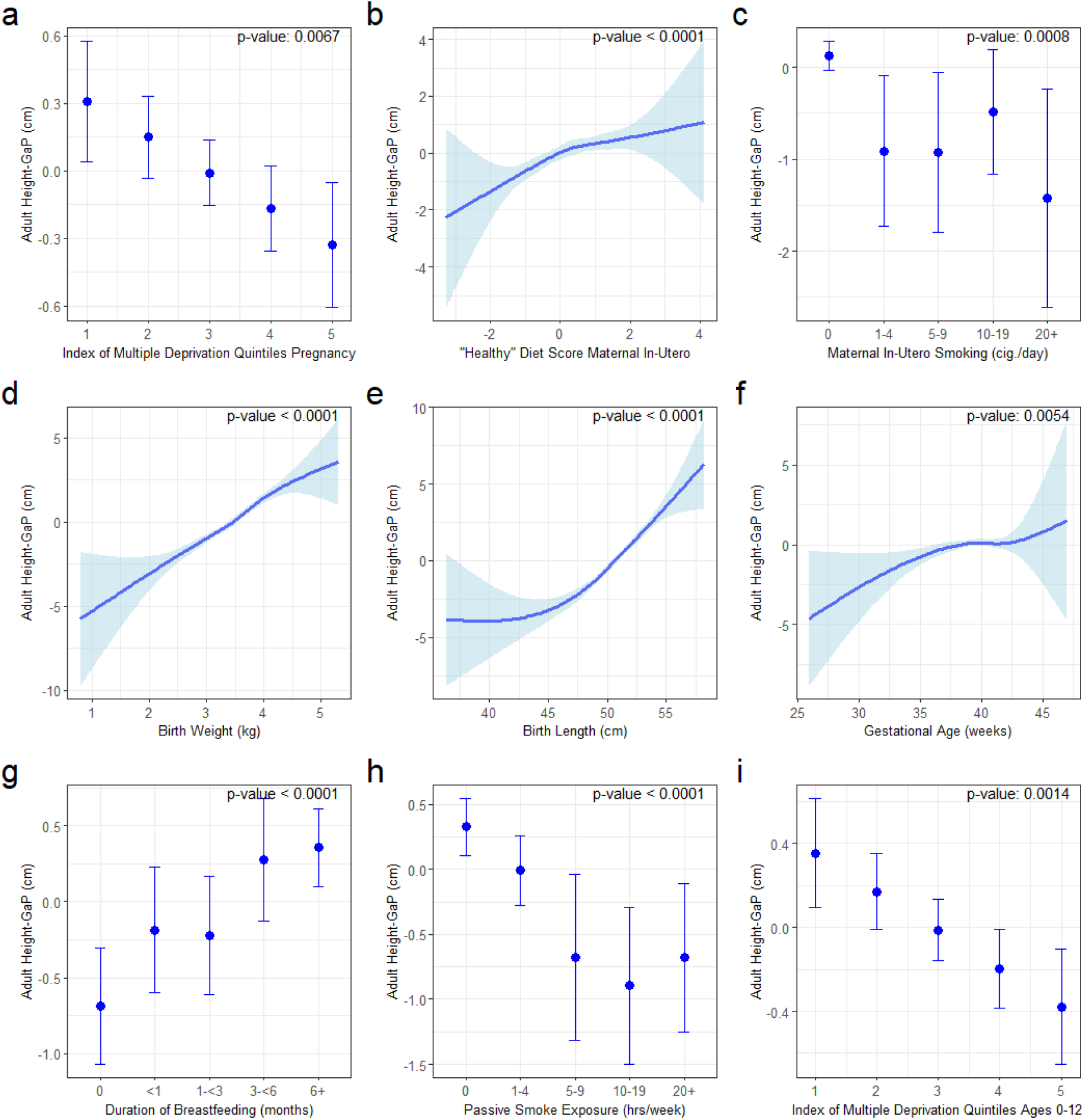
Estimated marginal mean adult height-GaP and 95%CI associated with early-life growth conditions in ALSPAC. Models adjusted for sex, age at height-GaP assessment, and genotype-predicted height. a) maternal English Index of Multiple Deprivation during pregnancy, b) maternal “healthy” diet score during pregnancy, c) maternal smoking during pregnancy, d) birth weight, e) birth length, f) gestational age, g) duration of breastfeeding, h) environmental tobacco smoke exposure at postnatal ages 6-54 months, i) child mean Index of Multiple Deprivation ages 0-12. Abbreviations: ALSPAC = Avon Longitudinal Study of Parents and Children; CI = confidence interval.

In the main adjusted model, larger adult height-GaP deficits were observed among participants with greater levels of multiple deprivation during pregnancy (mean height-GaP difference per quintile of multiple deprivation: -0.16 cm; 95%CI: -0.28 to -0.05 cm; p=0.006), greater maternal smoking during pregnancy (mean height-GaP difference comparing 0 vs. 20+ cigarettes per day: -1.51 cm; 95%CI: -2.71 to -0.30 cm; p=0.001), lower maternal “healthy diet” principal component during pregnancy (mean height-GaP difference per 1-SD decrement in “healthy diet”: -0.43 cm; 95%CI: -0.59 to -0.27 cm; p<0.001), lower gestational age at birth (mean height-GaP difference comparing <32 weeks to 38+ week: -4.09 cm; 95%CI: -7.14 to - 1.03 cm; p=0.005), lower birth weight (mean height-GaP difference per 1-kg decrement: -2.22 cm; 95%CI: -2.63 to -1.82 cm; p<0.001), lower birth length (mean height-GaP difference per 1-cm decrement: -0.67 cm; 95%CI: -0.78 to -0.55 cm; p<0.001), less breastfeeding (mean height-GaP difference comparing 0 vs. 6+ months: -1.03 cm; 95%CI: -1.50 to -0.56 cm; p=0.001), higher levels of multiple deprivation in childhood (mean height-GaP difference per quintile of multiple deprivation: -0.22 cm; 95%CI: -0.34 to -0.09 cm; p=0.001), greater household tobacco smoke exposure during childhood (mean height-GaP difference comparing 0 vs. 20+ hours/week -1.03 cm; 95%CI: -1.64 to -0.42 cm; p<0.001). Adjusted associations of adult height-GaP with residential outdoor PM_2.5_ exposure in the first year of life (p=0.593) and “healthy diet” principal component at 3 years old (p=0.026) did not meet the Bonferroni-corrected threshold of statistical significance. The variance in adult height-GaP explained by a multi-variable regression model including the aforementioned early-life growth conditions was 12.1% (95%CI: 9.8-14.5%).

Unadjusted associations of adult height-GaP with early-life growth conditions and the associations in DMHDS were consistent (*Supplemental Figure 2 and Tables 7-8*).

### Adult height-GaP and later-life mortality

The associations of adult height-GaP with mortality in UKBiobank are summarized in ***Figure 2*** and ***Table 3***.

**Figure 2:**
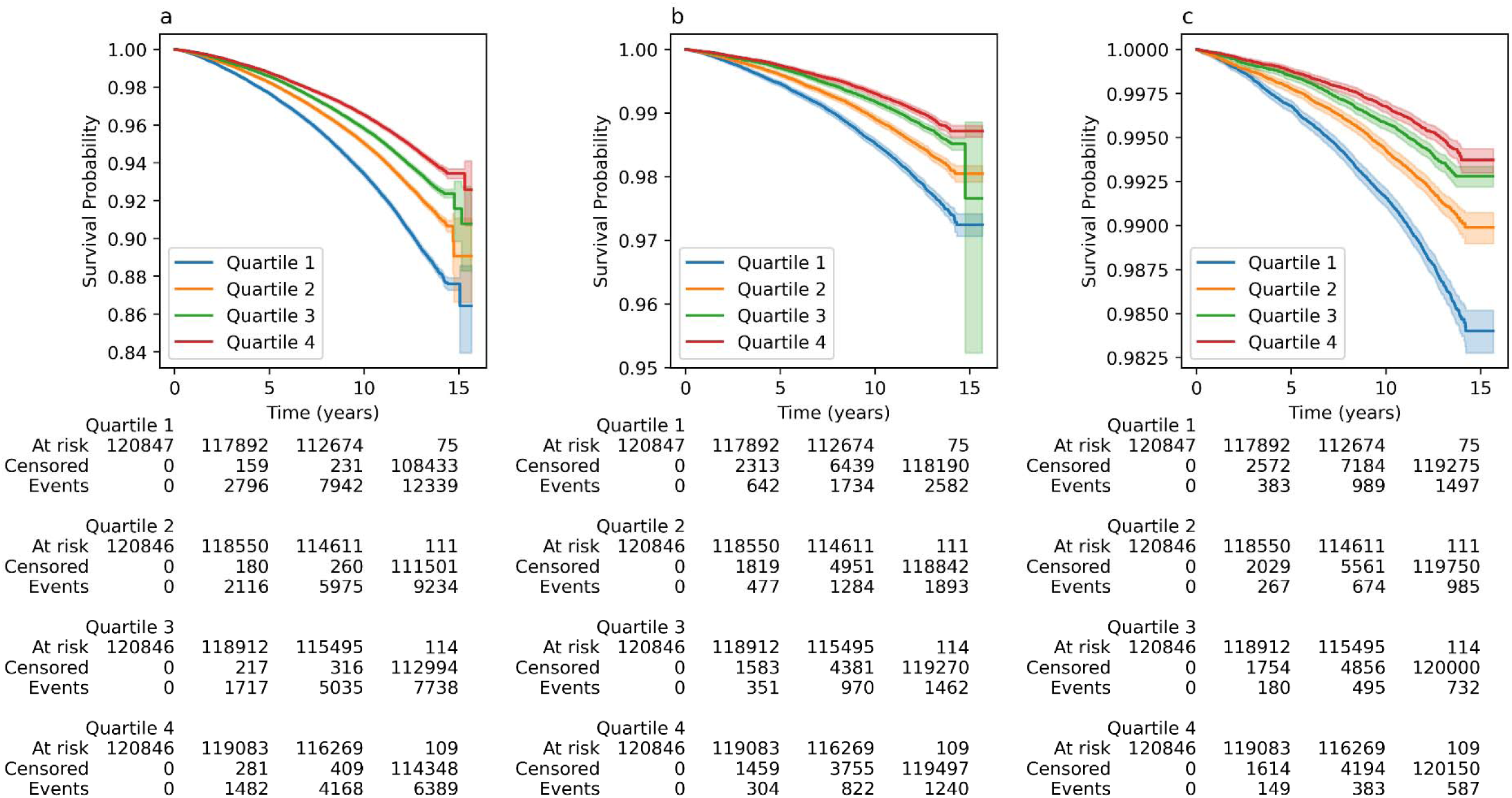
Kaplan-Meier survival curves by height-GaP quartile for a) all-cause mortality, b) atherosclerotic cardiovascular disease mortality and c) coronary heart disease mortality in the UKBiobank. Lower height-GaP quartile represents larger height-GaP deficit.

**Table 3:** Association of height-GaP with mortality in UKBiobank and MESA.

|  | **Hazard ratio per 1-SD height-GaP deficit (95% CI) p-value** |  |
| --- | --- | --- |
|  | **UKBiobank** | **MESA** |
| All-cause death |  |  |
| Model 1 | 1.11 (1.10, 1.13) p<0.001 | 1.09 (1.03, 1.15) p=0.004 |
| Model 2 | 1.12 (1.10, 1.13) p<0.001 | 1.09 (1.03, 1.15) p=0.004 |
| Atherosclerotic cardiovascular disease death |  |  |
| Model 1 | 1.15 (1.13, 1.18) p<0.001 | 1.17 (1.02, 1.34) p=0.027 |
| Model 2 | 1.15 (1.13, 1.18) p<0.001 | 1.17 (1.02, 1.34) p=0.027 |
| Atherosclerotic coronary heart disease death |  |  |
| Model 1 | 1.25 (1.22, 1.29) p<0.001 | 1.35 (1.14, 1.59) p<0.001 |
| Model 2 | 1.25 (1.22, 1.29) p<0.001 | 1.35 (1.14, 1.59) p<0.001 |

Model 1: age, sex, principal components of genetic ancestry.
Model 2: Model 1 variables, genotype-predicted height.
Abbreviations: MESA = Multi-Ethnic Study of Atherosclerosis.

In the main adjusted model, a 1-SD deficit in adult height-GaP (-5.2 cm) was associated with higher all-cause mortality (hazard ratio: 1.12; 95%CI: 1.10-1.13). A 1-SD deficit in adult height-GaP was also associated with a higher mortality from atherosclerotic cardiovascular disease (hazard ratio: 1.15; 95%CI: 1.13-1.18) and coronary heart disease (hazard ratio: 1.25; 95%CI: 1.22-1.29). Height-GaP deficit associations with mortality (all-cause, atherosclerotic cardiovascular, and coronary heart disease) were consistent in MESA (**Table 3**). There was no evidence of heterogeneity by sex- or by race-ethnicity in UKBiobank or MESA (p-interaction>0.231). Genotype-predicted height was not associated with higher all-cause or atherosclerotic cardiovascular disease mortality in either UKBiobank or MESA. Measured height—reflecting the linear combination of genotype-predicted height and height-GaP—was associated with both outcomes (*Supplemental Table 9*).

### Sensitivity analyses

Accounting for potential height-GaP-mortality confounding by later-life height loss using the median and worst-case scenario annualized height loss to “correct” measured height (and thus height-GaP) did not alter the statistical significance of mortality associations and only minimally attenuated the magnitude of association estimates (*Supplemental Table 10*). Additional adjustment for adult health determinants did not alter statistical significance and minimally attenuated mortality association estimates (*Supplemental Table 11*). Associations were consistent when restricting the ALSPAC sample to those with non-missing data or using ancestry-specific polygenic height scores (*Supplemental Tables 12-13)*.

## DISCUSSION

This study provides robust evidence that the difference between measured and genotype-predicted adult height—here termed “height-GaP”— can serve as a quantitative index of early-life growth adversity and as a predictor of later-life health. In multiple well-characterized cohorts, we demonstrate that adult height-GaP deficit is associated with prospectively ascertained adverse early-life events and exposures—including earlier gestational age at birth, less breastfeeding, greater household tobacco smoke exposure and socioeconomic deprivation— and is also associated with later-life mortality, including death from atherosclerotic cardiovascular disease. These findings support height-GaP as a simple composite index of early-life growth adversity that can be assessed in adulthood, thereby circumventing many of the challenges inherent to the investigation of developmental origins of health and disease.

We observed a clear gradient between adversity during the period of ontogenetic growth and subsequent height-GaP deficit in adulthood in two birth cohorts. These observations are consistent with literature documenting that noxious exposures, suboptimal nutritional environments and socioeconomic disadvantage during fetal and postnatal life attenuate children’s realized height.^28–30^ The variance in adult height-GaP explained by early-life growth conditions in ALSPAC was 12.1%. Although this fraction may appear modest, it is striking given that we relied on single or intermittent assessments of complex exposures (e.g., diet, neighborhood socioeconomic conditions) that likely exert dynamic influences across multiple developmental stages. Height-GaP will facilitate future research seeking to identify the periods of growth susceptibility and emerging threats to healthy human development.

We observed consistent associations between height-GaP and mortality in two adult cohorts including death from atherosclerotic cardiovascular disease—an important observation given that suboptimal early-life growth has long been linked to cardiovascular health in adulthood.^3–7^ The lack of association of genotype-predicted height with mortality underscores the potential precision of height-GaP to serve as a surrogate endpoint in early-life intervention trials seeking to improve lifelong health.^31^

The biological mechanisms underlying the early-life and later-life height-GaP associations are complex and were not directly interrogated in this study. The same prenatal and early-childhood exposures that impair growth—for instance, poor nutrition, socioeconomic adversity, or exposure to pollutants—may have lasting “programming” effects on immune, metabolic or other homeostatic systems, which in turn, may augment susceptibility to subclinical insults across the life course that culminate in premature mortality. Further mechanistic research should explore whether height-GaP correlates with dysregulation of such systems, thereby elucidating targetable links between early-life adversity and later-life health.

The findings of this study should be interpreted in the context of its limitations. First, the polygenic height score was derived from a sample composed predominantly of European ancestry individuals, which may limit generalizability to other ancestries or regions. We note, however, that the polygenic height score derivation sample included several other ancestries (East Asian: 472,730, Hispanic: 455,180, African: 293,593, South Asian: 77,890).^2^ Moreover, later-life mortality associations were homogeneous across race-ethnic groups in the Multi-Ethnic Study of Atherosclerosis. Nevertheless, genetic maps of height and validation of height-GaP associations across more diverse ancestries and geographies are needed. Second, GWAS-derived variant effect estimates may reflect both direct and indirect genetic effects. We note, however, that indirect genetic effects included in the polygenic height score would tend to attenuate associations between early life growth conditions and adult height-GaP. Third, the polygenic height score excludes rare variants. We do not believe this would substantively impact height-GaP applications because i) rare variants with weak height effects would have weak impact on an individual’s height-GaP, and ii) rare variants with large height effects (e.g., genetic skeletal dysplasias) would be clinically apparent. Fourth, height-GaP associations with later-life health outcomes may reflect residual confounding or mediation by adult health behaviours that correlate with adverse early-life growth conditions.^32^ We believe this is less likely because of consistency of findings in sensitivity analyses that adjusted for adult risk factors.

## CONCLUSION

This study introduces a novel quantitative index of early-life growth adversity, readily deployable in adulthood, to advance understanding of the developmental origins of disease and facilitate efforts to improve health across the life course.

## Data Availability

Due to legal and privacy reasons, participant-level data that support the findings of the study are managed under restricted access and available upon reasonable request from the ALSPAC, Dunedin, UKBiobank and MESA cohort study committees.

https://www.bristol.ac.uk/alspac/

https://dunedinstudy.otago.ac.nz/for-investigators/policy-statement-code-of-practic

https://www.ukbiobank.ac.uk/

https://internal.mesa-nhlbi.org/about

## ACKNOWLEDGEMENTS

This research was supported in part by grants from Canadian Institutes of Health Research (193740) and the U.S. National Institutes of Health (R01-HL130506, R01-HL077612).

The UK Medical Research Council and Wellcome (Grant ref: 217065/Z/19/Z) and the University of Bristol provide core support for ALSPAC. This publication is the work of the authors and will serve as guarantors for the contents of this paper. Genome-wide genotyping data was generated by Sample Logistics and Genotyping Facilities at Wellcome Sanger Institute and LabCorp (Laboratory Corporation of America) using support from 23andMe. A comprehensive list of grants funding and data access policy is available on the ALSPAC website: https://www.bristol.ac.uk/alspac/ We are extremely grateful to all the families who took part in this study, the midwives for their help in recruiting them, and the whole ALSPAC team, which includes interviewers, computer and laboratory technicians, clerical workers, research scientists, volunteers, managers, receptionists and nurses.

The UKBiobank data was accessed using the UK Biobank Resource under application number 773778. A comprehensive list of grant funding and data access policy is available on the UKBiobank website: https://www.ukbiobank.ac.uk/

MESA was supported by contracts 75N92020D00001, HHSN268201500003I, N01-HC-95159, 75N92020D00005, N01-HC-95160, 75N92020D00002, N01-HC-95161, 75N92020D00003, N01-HC-95162, 75N92020D00006, N01-HC-95163, 75N92020D00004, N01-HC-95164, 75N92020D00007, N01-HC-95165, N01-HC-95166, N01-HC-95167, N01-HC-95168 and N01-HC-95169 from the National Heart, Lung, and Blood Institute, and by grants UL1-TR-000040, UL1-TR-001079, and UL1-TR-001420 from the National Center for Advancing Translational Sciences (NCATS). Whole genome sequencing (WGS) for the Trans-Omics in Precision Medicine (TOPMed) program was supported by the National Heart, Lung and Blood Institute (NHLBI). WGS for “NHLBI TOPMed: Multi-Ethnic Study of Atherosclerosis (MESA)” (phs001416.v1.p1) was performed at the Broad Institute of MIT and Harvard (3U54HG003067-13S1). Centralized read mapping and genotype calling, along with variant quality metrics and filtering were provided by the TOPMed Informatics Research Center (3R01HL-117626-02S1). Phenotype harmonization, data management, sample-identity QC, and general study coordination, were provided by the TOPMed Data Coordinating Center (3R01HL-120393-02S1). The information contained herein was derived in part from data provided by the Bureau of Vital Statistics, New York City Department of Health and Mental Hygiene. The authors thank the other investigators, the staff, and the participants of the MESA study for their valuable contributions. A full list of participating MESA investigators and institutions and data access policy can be found at http://www.mesa-nhlbi.org.

We thank the Dunedin Study members, their families, and friends for their long-term involvement. The Dunedin Multidisciplinary Health and Development Research Unit is based at University of Otago within the Nga i Tahu tribal area whom we acknowledge as first peoples, tangata whenua (transl. people of this land). We thank Dunedin Unit research staff, previous Study Director, Emeritus Distinguished Professor, the late Richie Poulton, for his leadership during the Study’s research transition from young adulthood to aging (2000-2023) and Study founder, Dr Phil A. Silva. The Dunedin Multidisciplinary Health and Development Research Unit is supported by the New Zealand Health Research Council (Programme Grant 16-604) and has also received funding from the New Zealand Ministry of Business, Innovation and Employment. The study is also supported by the US-National Institute of Aging Grant R01AG032282, and UK Medical Research Council Grant MR/X021149/1. The data access policy can be found at https://dunedinstudy.otago.ac.nz/.

