## Supplemental Methods, Figures and Tables for "Quantifying the impact of early-life growth adversity on later-life health"

**SUPPLEMENT**

**Contents:**

**Supplemental Methods**	**2**

**Supplemental Figures and Tables**	**3**

Figures	3

Tables	5

**Supplemental Methods:**

Later-life height loss sensitivity analyses: To evaluate later-life height loss as a potential confounder of the later-life height-GaP mortality associations, we first estimated the sex-specific age at which height loss was first evident using longitudinal height measurements in UKBiobank. Next, we estimated the age- and sex-specific percentile distribution of annualized rate of height loss from the age of onset using quantile regression. Finally, we corrected each participant’s measured height (and thus height-GaP) back to the age of height loss onset using i) the median annualized height loss and the participant’s age and sex, and ii) a ‘worst case’ scenario of height-loss confounding. Under the ‘worst case’ scenario, on a percentile-by-percentile basis, a participant with larger height-GaP deficit was assumed to have experienced greater later-life height loss. For example, participants in the 95th percentile of height-GaP deficit are assumed to have experienced the 95th percentile of annualized height loss, and their measured height (and thus height-GaP) are corrected accordingly. This procedure is repeated on a percentile-by-percentile basis. Height loss-adjusted height-GaP values were then used to compute mortality associations adjusting for the same covariables listed above.

**Supplemental Figure and Tables:**

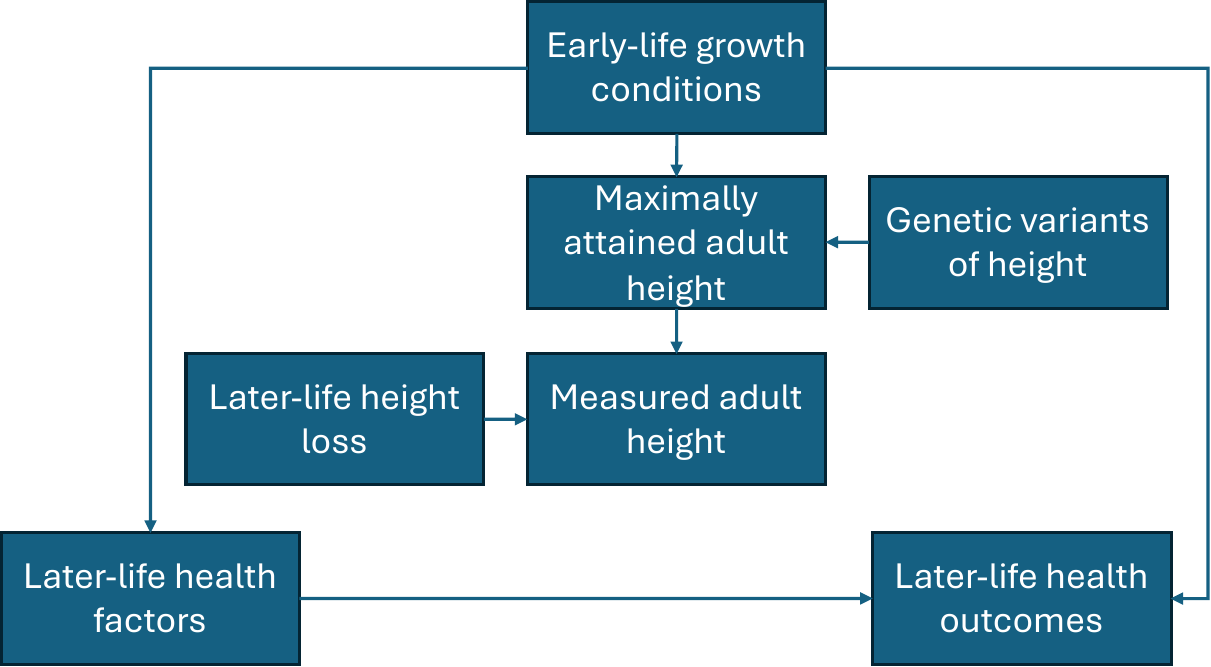

*Supplemental Figure 1*. A directed acyclic graph depicting hypothesized causal relationships of early-life growth conditions with late-life health outcomes.

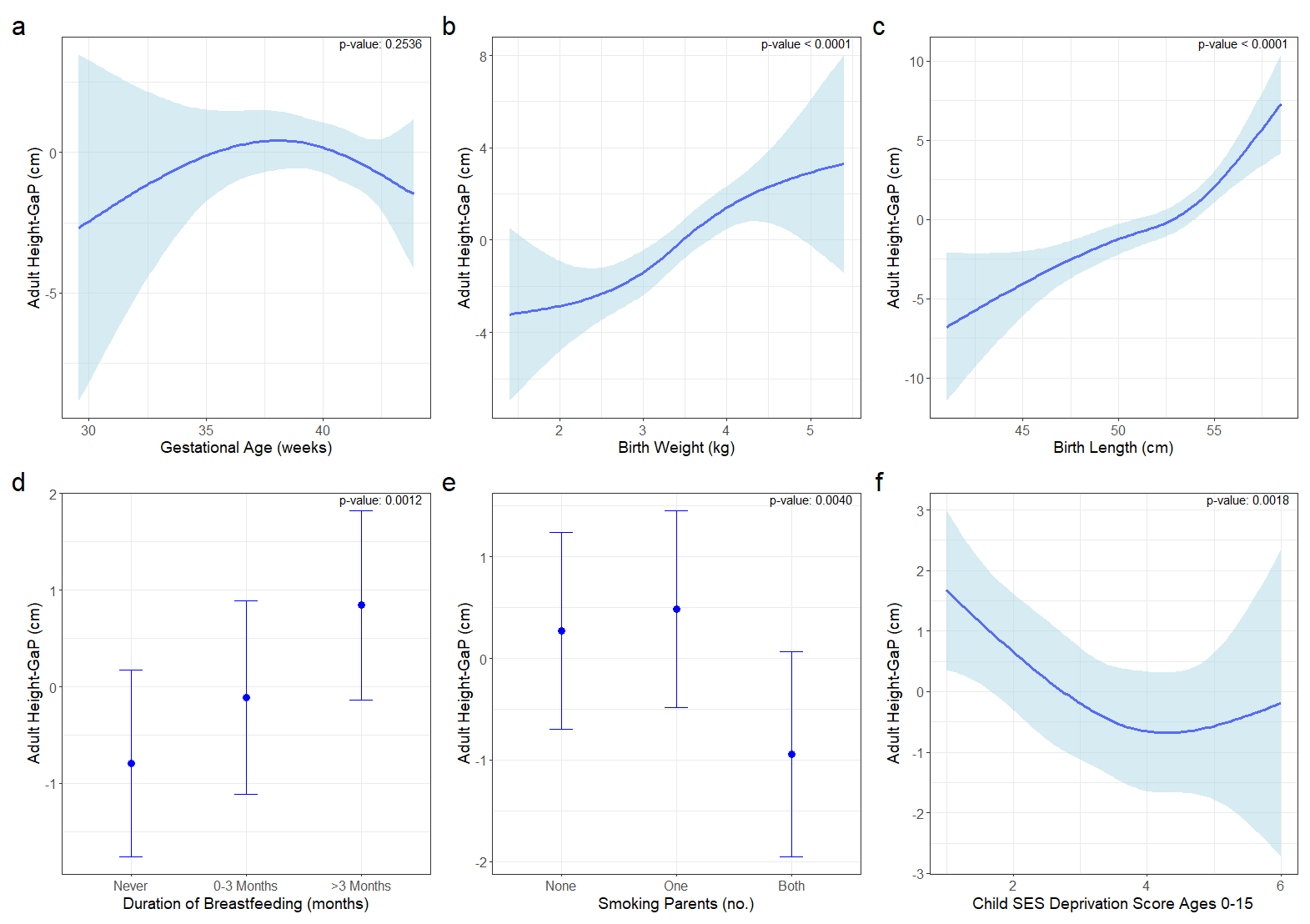

*Supplemental Figure 2*: Estimated marginal mean adult height-GaP and 95%CI associated with early-life growth conditions assessed in the Dunedin Multidisciplinary Health and Development Study. Models were adjusted for sex, age at height-GaP assessment, and genotype-predicted height. a) gestational age at birth, b) birth weight, c) birth length, d) duration of breastfeeding, e) number of tobacco-smoking parents, f) socio-economic status in quintiles between ages 0-15.

Abbreviations: SES = socio-economic status

*Supplemental Table 1:* Definitions and units of variables included in this study by cohort:

| **Variable** | **Cohort** | | | |
| --- | --- | --- | --- | --- |
|  | **ALSPAC** | **DMHDS** | **UKBiobank** | **MESA** |
| Age | Date of birth to study visit, years (integer) | Date of birth to study visit, years (integer) | Date of birth to study visit, years (integer) | Self-reported date of birth to study visit, years (integer) |
| Sex | Self-report, fixed category | Documented at birth, fixed category | Self-report, fixed category | Self-report, fixed category: male, female |
| Height | Standing shoeless, cm, integer | Standing shoeless, cm, integer | Standing shoeless, cm, integer | Standing shoeless, cm, integer |
| Adult weight | NA | NA | Standing bare foot on a Tanita BC418MA body composition analyzer | Standing shoeless, in light clothes with pockets emptied and jewelry removed, kg |
| Adult body mass index class | NA | NA | BMI category labelling based on continuous measurements:  <18.5 kg/m^2^  18.5 to <25 kg/m^2^  25 to <30 kg/m^2^  30+ kg/m^2^ | <18.5 kg/m^2^  18.5 to <25 kg/m^2^  25 to <30 kg/m^2^  30+ kg/m^2^ |
| Gestational age at birth | Time from date of mother’s last menstrual period to birth, weeks, based on mother’s self-report or clinical ultrasound assessment | Calculated from the date of last menstrual period when this was recalled with confidence. | NA | NA |
| Birth weight | Derived from birthweight data from obstetric data, ALSPAC measures and from birth notification/obstetric data, kilograms:  1. Value taken if identical from all sources  2. Lower value taken if disagreement between data sources <100g | Measured at birth, kilograms | NA | NA |
| Birth length | Crown-heel length using ALSPAC measures, cm | Measured at birth, mm. | NA | NA |
| Breastfeeding status and duration | Child-based questionnaire at 6 months of age, duration of breast feeding, fixed category:  1. <1 month  2. 1-<3 months  3. 3-<6 months  4. 6 or more  5. Missing | Number of weeks of breastfeeding reported by mother at the age 3 years assessment and validated from visiting nurse records | NA | NA |
| Index of multiple deprivation during pregnancy | Deprivation index in quintiles:  1 = low deprivated  2 = medium-low deprivated  3 = medium deprivated  4 = medium-high deprivated  5 = high deprivated | NA | NA | NA |
| Index of multiple deprivation during childhood | Mean deprivation index from birth to age 12 years. Higher quintiles indicate higher levels of deprivation. | Socioeconomic status based on the highest parental occupation recorded at each assessment between birth and age 15 years. Occupations were graded from 1 (high) to 6 (low) based on the income and education associated with that occupation in the New Zealand census. | NA | NA |
| Maternal smoking during pregnancy | No. of times smoked per day in the last two weeks, mother-based questionnaire at 18w gestation, category:  1. 0  2. 1-4  3. 5-9  4. 10-19  6. 20+  7. Missing | NA | NA | NA |
| Maternal “healthy” diet principal component during pregnancy | Derived dietary pattern scores for mother at 32 weeks pregnancy, component ‘Healthy’, according to mother-based questionnaire at 38 weeks gestation | NA | NA | NA |
| Household tobacco smoke during childhood | Estimated second-hand exposure to tobacco smoke, hours/week, from ages 6 months to 4.5 years. Categories: 1. 0  2. 1-4  4. 5-9  5. 10-19  6. 20+  7. Missing | Either parent reported to smoke at the age 7, 9, 11, or 13 year-old assessments. | NA | NA |
| “Healthy” diet principal component at 38 months | Derived dietary pattern scores at 38m age, component ‘Healthy’, according to child-based questionnaire at 3y2m of age | NA | NA | NA |
| Residential outdoor PM_2.5_ concentration | Particulate Matter <2.5 μg exposure average value at pregnancy, back-extrapolated using ratio method, μg/m^3^ | NA | NA | NA |
| Educational attainment | NA | NA | Self-reported, fixed category, multi-selection:  1. College or University degree  2. A levels/AS levels or equivalent  3. O levels/GCSEs or equivalent  4. CSEs or equivalent  5. NVQ or HND or HNC or equivalent  6. Other professional qualifications eg: nursing, teaching  7. None of the above  8. Prefer not to answer | Self-reported, fixed category:  1. No schooling  2. Grades 1-8  3. Grades 9-11  4. Completed high school or general educational development  5. Some college but no degree  6. Technical school certificate  7. Associate degrees  8. Bachelor’s degree  9. Graduate or professional degree |
| Family income, past 12 months | NA | NA | Self-reported, pre-tax, fixed category (£Pounds):  1. Less than 18,000  2. 18,000 to 30,999  3. 31,000 to 51,999  4. 52,000 to 100,000  5. Greater than 100,000  6. Do not know  7. Prefer not to answer | Self-reported, fixed category ($USD):  1: < $5,000  2: $5,000-7,999  3: $8,000-11,999  4: $12,000-15,999  5: $16,000-19,999  6: $20,000-24,999  7: $25,000-29,999  8: $30,000-34,999  9: $35,000-39,999  10: $40,000-49,999  11: $50,000-74,999  12: $75,000-99,999  13: $100,000+ |
| Health insurance status | NA | NA | NA | Self-reported, fixed categories:  1. Private  2. Medicare  3. Medicare + private  4. Medicaid  5. Military / Veteran affairs  6. Other  7. None |
| Race-ethnicity | Self-reported, fixed categories | Self-reported, fixed categories | Self-reported, fixed categories | Self-reported, fixed categories:  1. White  2. Black  3. Hispanic/Latino  4. Chinese |
| Cigarette smoking status | NA | NA | Self-reported | Self-reported |
| Never | NA | NA | Self-reported absence of past or former smoking behaviour | <100 lifetime cigarettes smoked |
| Former | NA | NA | Self-reported past tobacco smoking ‘smoked occasionally’ or ‘Smoked on most or all days’ | >=100 lifetime cigarettes smoked and cigarette smoked >30 days ago |
| Current | NA | NA | Self-reported current smoking ‘on most or all days’ or ‘Only occasionally’ | Cigarette smoked within 30 days |
| Pack-years | NA | NA | Number of cigarettes smoked per day / 20 * number of years smoking  Individuals who gave up smoking for >6 months is adjusted: Number of cigarettes smoked per day / 20 * (number of years smoking – 0.5) | Average number of cigarettes smoked per day while smoking / 20 * number of years smoking |
| Alcohol drinking status | NA | NA | Self-reported never, previous, current | Self-reported never, former, current |
| Drinks per week | NA | NA | Self-reported, fixed categories:  1. Daily or almost daily  2. Three or four times a week  3. Once or twice a week  4. One to three times a month  5. Special occasions only  6. Never  7. Prefer not to answer | Average number of drinks per week when drinking |
| Diabetes status | NA | NA | Self-reported previous diagnosis, yes, no, do not know, prefer not to answer | Fasting glucose >=126 mg/dl or use of glucose lowering medications |
| Hypertension status | NA | NA | Systolic blood pressure >= 140 mmHg or diastolic blood pressure > 90 mmHg or previous diagnosis of cardiovascular disease | Systolic blood pressure >= 140 mmHg or diastolic blood pressure >= 90 mmHg or hypertension medication use |
| Systolic blood pressure | NA | NA | Automated reading, mmHg | Average of 2nd and 3rd Dinamap systolic blood pressure measurements, in mm Hg |
| Diastolic blood pressure | NA | NA | Automated reading, mmHg | Average of 2nd and 3rd Dinamap diastolic blood pressure measurements, in mm Hg |
| Low density lipoprotein cholesterol concentration | NA | NA | Measured from randomly selected EDTA plasma samples using high-throughput NMR-based metabolic profiling platform, mmol/l, continuous | Friedewald equation-derived low density lipoprotein cholesterol concentration from overnight fasting serum measured total cholesterol, high density lipoprotein cholesterol and triglyceride |
| Hypertension medication use | NA | NA | Self-reported | Medication inventory |
| Diabetes medication use | NA | NA | Self-reported | Medication inventory |
| Lipid lowering medication use | NA | NA | Self-reported | Medication inventory |
| Moderate physical activity | NA | NA | Derived according to IPAQ guidelines, minutes/week, continuous | Self-reported questionnaire (Typical Week Physical Activity Survey)-derived moderate physical activity metabolic equivalent-minutes per week |
| Vigorous physical activity | NA | NA | Derived according to IPAQ guidelines, minutes/week, continuous | Self-reported questionnaire (Typical Week Physical Activity Survey)-derived vigorous physical activity metabolic equivalent-minutes per week |
| Death status | NA | NA | Derived from primary cause of death (ICD 10) according to National Death Registries | Derived from interval household phone contact every 9-12 months and query to National Death Index. |
| Death attributed to atherosclerotic cardiovascular disease | NA | NA | Derived from primary cause of death (ICD 10) according to National Death Registries, categories include:  I20-25 Ischemic heart diseases  I60 Subarachnoid haemorrhage  I61 Intracerebral haemorrhage  I63 Cerebral infarction  I64 Stroke, not specified as haemorrhage or infarction | Determined via standardized adjudication that included paired cardiologist or neurologist review of abstracted medical records, with disagreements resolved by full committee review. |
| Time to death | NA | NA | Date of death – date of enrollment divided by 365.25 days | Date of death – date of enrollment divided by 365.25 days |

Data dictionaries can be accessed at: <http://www.bristol.ac.uk/alspac/researchers/our-data/> <https://biobank.ndph.ox.ac.uk/showcase/> <https://dunedinstudy.otago.ac.nz/> <https://www.mesa-nhlbi.org/>

Abbreviations: ALSPAC = Avon Longitudinal Study of Parents and Children; DMHDS = Dunedin Multi-disciplinary Health and Development Study; MESA = Multi-Ethnic Study of Atherosclerosis; PM_2.5_ = particulate matter with diameter less than 2.5 micrometres; NA = not applicable

|  | **Included** | **Excluded** |
| --- | --- | --- |
| No. | 4,582 | 11,063 |
| Sex, no. (%) |  |  |
| Female | 2,579 (56.3) | 1,716 (40.7) |
| Male | 2,003 (43.7) | 2,499 (59.3) |
| Age at height-GaP assessment, years [IQR]] | 24 (18, 25) | 24 (18, 25) |
| Genotype-predicted height cm | 171.9 (7.9) | 173.8 (7.7) |
| Height-GaP, cm | 0.00 (5.06) | NA |
| Height, cm | 171.9 (9.4) | 170.5 (9.1) |
| Pregnancy index of multiple deprivation quintile | 2.91 (1.38) | 3.40 (1.40) |
| Maternal smoking during pregnancy, no. (%) |  |  |
| Never | 3751 (87.3) | 6913 (77.0) |
| 1-4 cigarettes/day, n (%) | 144 (3.4) | 384 (4.3) |
| 5-9 cigarettes/day, n (%) | 126 (2.9) | 510 (5.7) |
| 10-19 cigarettes/day, n (%) | 209 (4.9) | 842 (9.4) |
| 20+ cigarettes/day, n (%) | 67 (1.6) | 333 (3.7) |
| “Healthy” diet principal component during pregnancy | 0.05 (0.99) | -0.03 (1.01) |
| Gestational age at birth, weeks | 39.46 (1.83) | 37.90 (6.41) |
| Birth length, cm | 50.61 (2.13) | 50.46 (2.24) |
| Birth weight, kg | 3.43 (0.53) | 3.36 (0.60) |
| Breastfeeding duration, no. (%) |  |  |
| Never | 641 (15.8) | 2115 (29.9) |
| <1 month | 580 (14.3) | 1227 (17.4) |
| 1-<3 months | 623 (15.4) | 1140 (16.1) |
| 3-<6 months | 601 (14.8) | 819 (11.6) |
| 6+ months | 1608 (39.7) | 1764 (25.0) |
| Childhood index of multiple deprivation ages 0-12 quintile, mean | 2.77 (1.26) | 3.33 (1.34) |
| Childhood household tobacco smoke exposure duration, no. (%) |  |  |
| 0 hours/week | 1813 (54.2) | 1736 (41.8) |
| >0-5 hours/week | 984 (29.4) | 1247 (30.0) |
| >5-10 hours/week | 177 (5.3) | 296 (7.1) |
| >10-20 hours/week | 189 (5.6) | 365 (8.8) |
| 20+ hours/week | 183 (5.5) | 514 (12.4) |
| “Healthy” diet principal component at 38 months | 0.05 (0.99) | -0.03 (1.01) |
| Residential outdoor PM_2.5_ concentration, μg/m^3^ | 13.26 (0.77) | 13.34 (0.71) |

*Supplemental Table 2*: Characteristics of ALSPAC participants included and excluded from analyses.

Abbreviations: ALSPAC = Avon Longitudinal Study of Parents and Children; PM_2.5_ = particulate matter with diameter less than 2.5 micrometres.

*Supplemental Table 3*: Characteristics of participants in the UKBiobank included and excluded from the analysis.

|  | **Included** | **Excluded** |
| --- | --- | --- |
| No. | 483,385 | 18,994 |
| Sex, no. (%) |  |  |
| Female | 262,338 (54.3) | 109,69 (57.7) |
| Male | 221,047 (45.7) | 8,025 (42.3) |
| Age, years | 56.5 (8.1) | 57.9 (8.9) |
| Genotype-predicted height cm | 168.5 (7.7) | 169.1 (7.7) [n=3,777] |
| Height-GaP, cm | 0.0 (5.2) | -2.3 (5.1) [n=2,339] |
| Measured height, cm | 168.5 (9.3) | 167.1 (9.2) [n=16,453] |
| Body mass index class, no. (%) | [n=482,855] | [n=16,417] |
| Underweight (<18.5 kg/m^2^) | 2,495 (0.5) | 131 (0.7) |
| Healthy weight (18.5 to <25 kg/m^2^) | 157,422 (32.6) | 5,048 (26.6) |
| Overweight (25 to <30 kg/m^2^) | 205,325 (42.5) | 6,722 (35.4) |
| Obese (30+ kg/m^2^) | 117,613 (24.3) | 4,516 (23.8) |
| Missing |  |  |
| Ethnicity, no. (%) | 472,582 (94.1) | 455,857 (94.3) |
| White | 455,857 (94.4) | 16,694 (90.0) |
| Black | 7,511 (1.5) | 547 (2.9) |
| South Asian | 9,175 (1.9) | 704 (3.8) |
| Chinese | 1491 (0.3) | 82 (0.4) |
| Other | 7101(1.5) | 408 (2.2) |
| Do not know | 200 (0.0) | 17 (0.1) |
| Prefer not to answer | 1562 (0.3) | 99 (0.5) |
| Educational attainment, no. (%) |  |  |
| College or university degree | 156,536 (32.4) | 4,574 (24.1) |
| A/AS levels or equivalent | 53,759 (11.1) | 1,544 (8.1) |
| O levels/GCSEs or equivalent | 102,096 (21.1) | 3,074 (16.2) |
| CSEs or equivalent | 26,125 (5.4) | 760 (4.0) |
| NVQ/HND/HNC or equivalent | 31,777 (6.6) | 945 (5.0) |
| Other professional qualification | 24,944 (5.2) | 855 (4.5) |
| None of the above | 82,021 (17.0) | 3,237 (17.0) |
| Prefer not to answer | 5,210 (1.1) | 280 (1.5) |
| Missing | 917 (0.2) | 3,725 (19.6) |
| Household income, no. (%) |  |  |
| <£18,000 | 93,339 (19.3) | 3,838 (20.2) |
| £18,000-30,999 | 104,922 (21.7) | 3,223 (17.0) |
| £31,000-51,999 | 107,897 (22.3) | 2,851 (15.0) |
| £52,000-100,000 | 84,265 (17.4) | 1,980 (10.4) |
| >£100,000 | 22,406 (4.6) | 517 (2.7) |
| Do not know | 20,391 (4.2) | 908 (4.8) |
| Prefer not to answer | 47,959 (9.9) | 1,869 (9.8) |
| Missing | 2,206 (0.5) | 3,808 (20.0) |
| Smoking status, no. (%) |  |  |
| Never smoker | 265,660 (55.0) | 10,643 (56.0) |
| Former smoker | 166,886 (34.5) | 6,228 (32.8) |
| Current smoker | 50,839 (10.5) | 2,123 (11.2) |
| Pack-years among ever smokers, median (IQR) | 19.0 (9.9,32.0) | 20.7 (10.8,34.5) |
| Alcohol use, no. (%) |  |  |
| Never | 21,160 (4.4) | 1,220 (6.4) |
| Former | 17266 (3.6) | 827 (4.4) |
| Current | 443,785 (91.8) | 16,467 (86.7) |
| Frequency of Weekly Alcohol Intake, no. (%) |  |  |
| Daily or almost daily | 98,257 (20.3) | 3,490 (18.4) |
| 3-4 times weekly | 111,717 (23.1) | 3,700 (19.5) |
| 1-2 times weekly | 124,644 (25.8) | 4,617 (24.3) |
| 1-3 times monthly | 53,776 (11.1) | 2,061 (10.9) |
| Special occasions only | 55,391 (11.5) | 2,599 (13.7) |
| Never | 38,566 (8.0) | 2,059 (10.8) |
| Prefer not to answer | 603 (0.1) | 546 (0.1) |
| Missing | 899 (0.2) | 488 (0.1) |
| Diabetes mellitus, no. (%) | 25,090 (5.2) | 1,305 (6.9) |
| Hypertension, no. (%) |  |  |
| Systolic blood pressure, mmHg | 139.7 (19.7) | 141.2 (20.8) |
| Low density lipoprotein cholesterol, mg/dL | 3.6 (0.9) | 3.5 (0.9) |
| Lipid lowering medication use, no. (%) |  |  |
| Moderate-to-vigorous physical activity, MET-min/week, median (IQR) |  |  |
| Time to death or censorship, years | 12.4 (1.8) | 12.4 (2.6) |
| Deaths, no. (%) | 35,703 (7.4) | 2,194 (11.6) |
| Atherosclerotic cardiovascular disease, no. (%) | 11,066 (2.3) | 710 (3.7) |
| Atherosclerotic coronary heart disease, no. (%) | 3,801 (0.8) | 240 (1.3) |

Abbreviations: GCSE = General Certificate of Secondary Education; CSE = Certificate of Secondary Education; NVQ = National Vocational Qualifications; HND = Higher National Diploma; HNC =Higher National Certificate; IQR = interquartile range; MET = metabolic equivalent.

*Supplemental Table 4*: Characteristics of participants included in the Dunedin Multidisciplinary Health and Development Study analyses.

|  | **All** | **By Height-GaP Quartile** | | | |
| --- | --- | --- | --- | --- | --- |
|  |  | **1** | **2** | **3** | **4** |
| No. | 855 | 214 | 214 | 214 | 213 |
| Sex, no. (%) |  |  |  |  |  |
| Female | 419 (49.0) | 111 (51.9) | 103 (48.1) | 92 (43.0) | 113 (53.1) |
| Male | 436 (51.0) | 103 (48.1) | 111 (51.9) | 122 (57.0) | 100 (46.9) |
| Age at height-GaP assessment, years | 26 (26, 26) | 26 (26, 26) | 26 (26, 26) | 26 (26, 26) | 26 (26, 26) |
| Genotype-predicted height cm | 171.8 (7.4) | 171.6 (7.3) | 171.7 (7.5) | 172.7 (7.4) | 171.2 (7.4) |
| Height-GaP, cm | 0.0 (5.3) | -6.5 (2.7) | -1.8 (1.0) | 1.59 (1.0) | 6.7 (3.2) |
| Height, cm | 171.8 (9.1) | 165.1 (7.7) | 169.9 (7.6) | 174.3 (7.4) | 177.9 (8.3) |
| Gestational age at birth, weeks | 40.0 (1.7) | 40.1 (1.8) | 40.1 (1.7) | 40.0 (1.5) | 39.9 (1.6) |
| Birth length, cm | 51.9 (2.3) | 51.1 (2.4) | 51.8 (2.3) | 52.1 (2.0) | 52.6 (2.2) |
| Birth weight, kg | 3.39 (0.52) | 3.23 (0.54) | 3.35 (0.51) | 3.44 (0.46) | 3.52 (0.51) |
| Breastfeeding duration, no. (%) |  |  |  |  |  |
| Never | 371 (43.5) | 108 (50.9) | 100 (46.7) | 86 (40.2) | 77 (36.2) |
| 0-3 Months | 254 (29.8) | 58 (27.4) | 66 (30.8) | 70 (32.7) | 60 (28.2) |
| >3 Months | 228 (26.7) | 46 (21.7) | 48 (22.4) | 58 (27.1) | 76 (35.7) |
| Childhood (ages 0-15 years) socio-economic deprivation score | 3.19 (1.11) | 3.37 (1.05) | 3.21 (1.12) | 3.18 (1.08) | 3.02 (1.18) |
| Parents who smoke tobacco, no. (%) |  |  |  |  |  |
| None | 297 (36.5) | 73 (35.6) | 70 (34.3) | 75 (36.9) | 79 (39.3) |
| One | 266 (32.7) | 59 (28.8) | 64 (31.4) | 66 (32.5) | 77 (38.3) |
| Both | 250 (30.8) | 73 (35.6) | 70 (34.3) | 62 (30.5) | 45 (22.4) |

Mean (SD) unless otherwise noted.

Abbreviations: SD = standard deviation.

*Supplementary Table 5:* Characteristics of participants included in the Multi-Ethnic Study of Atherosclerosis analyses.

|  | **All** | **By Height-GaP Quartile** | | | |
| --- | --- | --- | --- | --- | --- |
|  |  | **1** | **2** | **3** | **4** |
| No. | 6,352 | 1,588 | 1,588 | 1,588 | 1,588 |
| Sex, no. (%) |  |  |  |  |  |
| Female | 3,323 (52.3) | 831 (52.3) | 830 (52.3) | 832 (52.3) | 830 (52.3) |
| Male | 3,029 (47.7) | 757 (47.7) | 758 (47.7) | 756 (47.6) | 758 (47.7) |
| Age, years | 62.2 (10.3) | 66.7 (10.1) | 63.5 (10.1) | 60.8 (9.7) | 58.0 (9.0) |
| Genotype-Predicted Height cm | 166.4 (8.3) | 166.4 (8.1) | 166.4 (8.3) | 166.3 (8.3) | 166.4 (8.3) |
| Height-GaP, cm | 0.0 (5.7) | -7.2 (2.9) | -1.8 (1.1) | 1.7 (1.1) | 7.2 (3.1) |
| Measured height, cm | 166.4 (10.0) | 159.2 (8.5) | 164.6 (8.3) | 168.0 (8.4) | 173.6 (9.0) |
| Body mass index class, no. (%) |  |  |  |  |  |
| Underweight (<18.5 kg/m^2^) | 56 (0.9) | 16 (1.0) | 16 (1.0) | 9 (0.6) | 15 (0.9) |
| Healthy weight (18.5 to <25 kg/m^2^) | 1,784 (28.1) | 415 (26.1) | 448 (28.2) | 481 (30.3) | 440 (27.7) |
| Overweight (25 to <30 kg/m^2^) | 2,492 (39.2) | 644 (40.6) | 645 (40.6) | 593 (37.3) | 610 (38.4) |
| Obese (30+ kg/m^2^) | 2,020 (31.8) | 513 (32.3) | 479 (30.2) | 505 (31.8) | 523 (32.9) |
| Self-reported race-ethnicity, no. (%) |  |  |  |  |  |
| White | 2,486 (39.1) | 576 (36.3) | 657 (41.4) | 668 (42.1) | 585 (36.8) |
| Black | 1,660 (26.1) | 464 (29.2) | 379 (23.9) | 363 (22.9) | 454 (28.6) |
| Hispanic | 1,437 (22.6) | 384 (24.2) | 336 (21.2) | 340 (21.4) | 377 (23.7) |
| Chinese | 769 (12.1) | 164 (10.3) | 216 (13.6) | 217 (13.7) | 172 (10.8) |
| Educational attainment, no. (%) | [n=6,332] | [n=1,582] | [n=1,584] | [n=1,582] | [n=1,584] |
| Less than high school | 1,144 (18.1) | 398 (25.2) | 308 (19.4) | 229 (14.5) | 209 (13.2) |
| Completed high school | 2,188 (34.6) | 567 (35.8) | 551 (34.8) | 523 (33.1) | 547 (34.5) |
| Technical school certificate / Associate’s degree | 768 (12.1) | 182 (11.5) | 174 (11.0) | 209 (13.2) | 203 (12.8) |
| Bachelor’s degree | 1,106 (17.5) | 215 (13.6) | 279 (17.6) | 293 (18.5) | 319 (20.1) |
| Graduate or professional degree | 1,126 (17.8) | 220 (13.9) | 272 (17.2) | 328 (20.7) | 306 (19.3) |
| Household income, no. (%) | [n=6,113] | [n=1,500] | [n=1,533] | [n=1,536] | [n=1,544] |
| <$25,000 | 1,937 (31.7) | 641 (42.7) | 529 (34.5) | 402 (26.2) | 365 (23.6) |
| $25,000-49,999 | 1,763 (28.8) | 432 (28.8) | 437 (28.5) | 462 (30.1) | 432 (28.0) |
| $50,000-74,999 | 1,034 (16.9) | 195 (13.0) | 236 (15.4) | 292 (19.0) | 311 (20.1) |
| $75,000-99,999 | 562 (9.2) | 92 (6.1) | 134 (8.7) | 147 (9.6) | 189 (12.2) |
| $100,000+ | 817 (13.4) | 140 (9.3) | 197 (12.9) | 233 (15.2) | 247 (16.0) |
| Health insurance status, no. (%) |  |  |  |  |  |
| Private | 3,409 (53.7) | 635 (40.0) | 790 (49.8) | 923 (58.1) | 1,061 (66.8) |
| Medicare | 1,077 (17.0) | 391 (24.6) | 293 (18.5) | 221 (13.9) | 172 (10.8) |
| Medicare + private | 1,209 (19.0) | 387 (24.4) | 331 (20.8) | 277 (17.4) | 214 (13.5) |
| None | 557 (8.8) | 145 (9.1) | 142 (8.9) | 145 (9.1) | 125 (7.9) |
| Medicaid | 100 (1.6) | 30 (1.9) | 32 (2.0) | 22 (1.4) | 16 (1.0) |
| Smoking status, no. (%) | [n=6,333] | [n=1,583] | [n=1,584] | [n=1,582] | [n=1,584] |
| Never smoker | 3,189 (50.4) | 854 (54.0) | 773 (48.8) | 809 (51.1) | 753 (47.5) |
| Former smoker | 2,314 (36.5) | 554 (35.0) | 628 (39.7) | 561 (35.5) | 571 (36.1) |
| Current smoker | 830 (13.1) | 175 (11.1) | 183 (11.6) | 212 (13.4) | 260 (16.4) |
| Pack-years among ever smokers, median (IQR) | 16 (6, 32) | 16 (5, 34) | 17 (6, 34) | 15 (6, 32) | 15 (5, 29) |
| Alcohol use, no. (%) | [n=6306] | [n=1,574] | [n=1,581] | [n=1,575] | [n=1,576] |
| Never | 1,307 (20.7) | 375 (23.8) | 328 (20.8) | 309 (19.6) | 295 (18.7) |
| Former | 1,502 (23.8) | 403 (25.6) | 384 (24.3) | 372 (23.6) | 343 (21.8) |
| Current | 3,497 (55.5) | 796 (50.6) | 869 (55.0) | 894 (56.8) | 938 (59.5) |
| No. of alcohol drinks per week among ever drinkers, median (IQR) | 2 (0, 6) | 2 (0, 6) | 2 (0, 6) | 2 (0, 6) | 2 (0, 6) |
| Diabetes mellitus, no. (%) | 786 (12.4)  [n=6,337] | 231 (13.6)  [n=1,584] | 184 (9.5)  [n=1,584] | 193 (12.2)  [n=1,582] | 178 (11.2)  [n=1,587] |
| Hypertension, no. (%) | 2,839 (44.6) | 838 (52.8) | 713 (44.9) | 659 (41.5) | 622 (39.2) |
| Systolic blood pressure, mmHg (SD) | 126.5 (21.5) | 123.3 (19.4) | 125.7 (21.5) | 126.6 (21.2) | 130.3 (23.3) |
| LDL cholesterol, mg/dL | 117.3 (31.5) | 117.2 (31.5) | 118.8 (32.3) | 117.9 (31.3) | 115.1 (30.9) |
| Lipid lowering medication use, no. (%) | 1,018 (16.0) | 304 (19.1) | 259 (16.3) | 230 (14.5) | 225 (14.2) |
| Moderate-to-vigorous physical activity, MET-min/week, median (IQR) | 4,020 (1,973, 7,545) | 3,495 (1,650, 6,900) | 4,001 (1,875, 7,513) | 4,215 (2,040, 7,815) | 4,388 (2,250, 7984) |
| Time to death or censorship, years | 13.6 (3.1) | 12.9 (3.6) | 13.6 (3.1) | 13.8 (2.9) | 14.0 (2.8) |
| Deaths, no. (%) | 1,337 (21.0) | 498 (31.4) | 333 (21.0) | 278 (17.5) | 228 (14.4) |
| Atherosclerotic cardiovascular disease, no. (%) | 233 (3.7) | 97 (6.1) | 55 (3.5) | 43 (2.4) | 38 (2.4) |
| Atherosclerotic coronary heart disease, no. (%) | 155 (2.4) | 74 (4.7) | 30 (1.9) | 26 (1.6) | 25 (1.6) |

Levels of educational attainment (total: 8) and household income (total: 13) were retained for regression analyses but combined in the Table for brevity. Square parentheses indicate the number of participants with non-missing values and the remaining participants were included in analyses using a missing indicator variable.

Abbreviations: SD = standard deviation; IQR = interquartile range.

*Supplemental Table 6*: Adjusted associations of early-life growth conditions with adult height-GaP in ALSPAC.

|  | **Adjusted mean difference in height-GaP, cm (95%CI) p-value** |
| --- | --- |
| Pregnancy index of multiple deprivation, per 1-quintile increment | -0.16 (-0.28, -0.05) p=0.006 |
| Maternal smoking during pregnancy | p=0.001 |
| Never | Reference |
| 1-4 cigarettes/day, n (%) | -1.03 (-1.86, -0.20) |
| 5-9 cigarettes/day, n (%) | -1.05 (-1.93, -0.17) |
| 10-19 cigarettes/day, n (%) | -0.61 (-1.31, 0.09) |
| 20+ cigarettes/day, n (%) | -1.51 (-2.71, -0.30) |
| “Healthy” diet principal component during pregnancy, per 1-SD decrement | -0.43 (-0.59, -0.27) p<0.001 |
| Gestational age at birth, weeks, no. (%) | p=0.005 |
| 38+ | Reference |
| 32-<38 | -0.08 (-0.59, 0.43) |
| <32 | -4.09 (-7.14, -1.03) |
| Birth length, per 1-cm decrement | -0.67 (-0.78, -0.55) p<0.001 |
| Birth weight, per 1-kg decrement | -2.22 (-2.63, -1.82) p<0.001 |
| Breastfeeding duration | p<0.001 |
| 6+ months | Reference |
| 3-<6 months | -0.06 (-0.53, 0.42) |
| 1-<3 months | -0.56 (-1.04, -0.08) |
| <1 month | -0.55 (-1.05, -0.05) |
| Never | -1.03 (-1.50, -0.56) |
| Childhood (ages 0-12 years) index of multiple deprivation, per 1-quintile increment | -0.22 (-0.34, -0.09) p=0.001 |
| Childhood (ages 6-34 months) household tobacco smoke exposure duration | p<0.001 |
| 0 hours/week | Reference |
| >0-5 hours/week | -0.34 (-0.70, 0.01) |
| >5-10 hours/week | -0.99 (-1.66, -0.31) |
| >10-20 hours/week | -1.22 (-1.86, -0.57) |
| 20+ hours/week | -1.03 (-1.64, -0.42) |
| “Healthy” diet principal component at 38 months, per 1-SD decrement | -0.22 (-0.41, -0.03) p=0.026 |
| Residential outdoor PM_2.5_ concentration, per 1-μg/m^3^ increment | 0.06 (-0.15,0.26) p=0.593 |

Model covariables: age at height-GaP assessment, sex, genotype-predicted height.

Abbreviations: ALSPAC = Avon Longitudinal Study of Parents and Children; SD = standard deviation; PM_2.5_ = particulate matter with diameter less the 2.5 micrometres.

*Supplemental Table 7*: Unadjusted associations of early-life growth conditions with adult height-GaP in ALSPAC.

|  | **Unadjusted mean difference in height-GaP, cm (95%CI)** |
| --- | --- |
| Pregnancy index of multiple deprivation, per 1-quintile increment | -0.19 (-0.31, -0.08) p=0.001 |
| Maternal smoking during pregnancy | p=0.001 |
| Never | Reference |
| 1-4 cigarettes/day, n (%) | -1.06 (-1.90, -0.23) |
| 5-9 cigarettes/day, n (%) | -1.09 (-1.98, -0.21) |
| 10-19 cigarettes/day, n (%) | -0.64 (-1.34, 0.06) |
| 20+ cigarettes/day, n (%) | -1.54 (-2.75, -0.33) |
| “Healthy” diet principal component during pregnancy, per 1-SD decrement | -0.46 (-0.62, -0.30) p<0.001 |
| Gestational age at birth, weeks, no. (%) | p=0.005 |
| 38+ | Reference |
| 32-<38 | -0.07 (-0.58, 0.44) |
| <32 | -4.14 (-7.22, -1.05) |
| Birth length, per 1-cm decrement | -0.62 (-0.72, -0.51) p<0.001 |
| Birth weight, per 1-kg decrement | -2.16 (-2.55, -1.77) p<0.001 |
| Breastfeeding duration | p<0.001 |
| 6+ months | Reference |
| 3-<6 months | -0.06 (-0.54, 0.42) |
| 1-<3 months | -0.60 (-1.08, -0.12) |
| <1 month | -0.62 (-1.12, -0.11) |
| Never | -1.12 (-1.59, -0.64) |
| Childhood (ages 0-12 years) index of multiple deprivation, per 1-quintile increment | -0.25 (-0.38, -0.13) p<0.001 |
| Childhood (ages 6-34 months) household tobacco smoke exposure duration | p<0.001 |
| 0 hours/week | Reference |
| >0-5 hours/week | -0.40 (-0.75, -0.04) |
| >5-10 hours/week | -1.08 (-1.76, -0.40) |
| >10-20 hours/week | -1.31 (-1.96, -0.67) |
| 20+ hours/week | -1.13 (-1.74, -0.51) |
| “Healthy” diet principal component at 38 months, per 1-SD decrement | -0.21 (-0.40, -0.01) |
| Residential outdoor PM_2.5_ concentration, per 1-μg/m^3^ increment | 0.06 (-0.15, 0.26) p=0.595 |

Abbreviations: ALSPAC = Avon Longitudinal Study of Parents and Children; SD = standard deviation; PM_2.5_ = particulate matter with diameter less the 2.5 micrometres.

*Supplemental Table 8*: Adjusted associations of early-life growth conditions with adult height-GaP in the Dunedin Multidisciplinary Health and Development Study.

|  | **Adjusted mean difference in height-GaP cm (95%CI) p-value** |
| --- | --- |
| Maternal smoking during pregnancy | p=0.239 |
| No | Reference |
| Yes | -0.46 (-1.22, 0.30) |
| Gestational age at birth, weeks, no. (%) | p=0.347 |
| 38+ | Reference |
| 32-<38 | 0.47 (-0.82, 1.75) |
| <32 | -6.62 (-17.01, 3.78) |
| Birth length, cm, per 1-cm decrement | -0.63 (-0.78, -0.47) p<0.001 |
| Birth weight, kg, per 1-kg decrement | -2.43 (-3.11, 1.75) p<0.001 |
| Breastfeeding duration | p=0.001 |
| 3+ months | Reference |
| 0-<3 months | -0.96 (-1.90, -0.01) |
| Never | -1.64 (-2.51, -0.77) |
| Childhood (ages 0-15 years) index of socio-economic deprivation, per 1-SD increment | -0.63 (-0.98, -0.28) p=0.001 |
| Tobacco-smoking parents | p=0.004 |
| None | Reference |
| One | -0.96 (-1.90, -0.01) |
| Both | -1.64 (-2.51, -0.77) |

Model covariables: age at height-GaP assessment, sex, genotype-predicted height.

Abbreviations: SD = standard deviation.

*Supplemental Table 9*: Association of measured height, genotype-predicted height and height-GaP with mortality in UKBiobank and MESA.

|  | **UKBiobank** | | | |  | **MESA** | | | |
| --- | --- | --- | --- | --- | --- | --- | --- | --- | --- |
|  | **Hazard ratio per 1-SD deficit in Measured height** |  | **Hazard ratio per 1-SD deficit in genotype-predicted height** | **Hazard ratio per 1-SD deficit in Height-GaP** |  | **Hazard ratio per 1-SD deficit in measured height** |  | **Hazard ratio per 1-SD deficit in genotype-predicted height** | **Hazard ratio per 1-SD deficit in height-GaP** |
| All-cause  death | 1.08  (1.06, 1.09) p<0.001 |  | 0.97  (0.96, 0.99)  p<0.001 | 1.12  (1.10, 1.13) p<0.001 |  | 1.08  (1.01, 1.15) p=0.016 |  | 1.00  (0.93, 1.08)  p=0.971 | 1.09  (1.03, 1.15)  p=0.004 |
| Atherosclerotic cardiovascular disease death | 1.12  (1.10, 1.15) p<0.001 |  | 1.00  (0.98, 1.03)  p=0.933 | 1.15  (1.13, 1.18) p<0.001 |  | 1.14  (0.98, 1.32) p=0.095 |  | 0.97  (0.82, 1.15)  p=0.740 | 1.17  (1.02, 1.34) p=0.027 |
| Atherosclerotic coronary heart disease death | 1.22  (1.18, 1.26) p<0.001 |  | 1.02  (0.98, 1.06)  p=0.282 | 1.25  (1.22, 1.29) p<0.001 |  | 1.31  (1.09, 1.57) p=0.005 |  | 0.98  (0.79, 1.21)  p=0.846 | 1.35  (1.14, 1.59) p<0.001 |

The models estimating the measured height associations with mortality included age, sex, principal components of genetic ancestry and measured height.

The models estimating the genotype-predicted height and height-GaP associations with mortality included age, sex and principal components of genetic ancestry, genotype-predicted height and height-GaP.

Abbreviations: MESA = Multi-Ethnic Study of Atherosclerosis.

*Supplemental Table 10*: Association of height-GaP with mortality in UKBiobank accounting for later-life height loss.

|  | **Hazard ratio per 1-SD height-GaP deficit (95%CI) p-value** | |
| --- | --- | --- |
|  | **Median height-loss corrected height-GaP** | **Worst case scenario height-loss corrected height-GaP** |
| All-cause death |  |  |
| Model 1 | 1.11 (1.10, 1.12) p<0.001 | 1.07 (1.06, 1.07) p<0.001 |
| Model 2 | 1.11 (1.10, 1.13) p<0.001 | 1.07 (1.06, 1.07) p<0.001 |
| Atherosclerotic cardiovascular disease death |  |  |
| Model 1 | 1.15 (1.12, 1.18) p<0.001 | 1.09 (1.07, 1.10) p<0.001 |
| Model 2 | 1.15 (1.12, 1.18) p<0.001 | 1.09 (1.07, 1.10) p<0.001 |
| Atherosclerotic coronary heart disease death |  |  |
| Model 1 | 1.25 (1.21, 1.29) p<0.001 | 1.14 (1.12, 1.17) p<0.001 |
| Model 2 | 1.25 (1.21, 1.29) p<0.001 | 1.14 (1.12, 1.17) p<0.001 |

Model 1: Age, sex, principal components of genetic ancestry.

Model 2: Model 1 variables, genotype-predicted height.

Abbreviations: SD = standard deviation.

*Supplemental Table 11*: Association of height-GaP with mortality adjusted for adult health conditions in UKBiobank and MESA.

|  | **Hazard ratio per 1-SD height-GaP deficit (95%CI) p-value** | |
| --- | --- | --- |
|  | **UKBiobank** | **MESA** |
| All-cause death | 1.05 (1.03, 1.06) p<0.001 | 1.09 (1.03, 1.16) p=0.006 |
| Atherosclerotic cardiovascular disease death | 1.06 (1.03, 1.09) p<0.001 | 1.18 (1.02, 1.36) p=0.022 |
| Atherosclerotic coronary heart disease death | 1.12 (1.09, 1.16) p<0.001 | 1.32 (1.10, 1.57) p=0.003 |

Model covariables: age, sex, principal components of genetic ancestry, genotype-predicted height, cigarette smoking status, pack-years, alcohol use status, drinks per week, minutes of moderate and of vigorous physical activity per week, weight class, diabetes status, hypertension status, systolic blood pressure, LDL cholesterol concentration, cholesterol-lowering medication use, educational attainment, health insurance status and household income.

*Supplementary Table 12*: Association of early-life growth conditions with adult height-GaP in ALSPAC restricted to participants with non-missing data.

|  | **Adjusted mean difference in height-GaP (95%CI) p-value** |
| --- | --- |
| Pregnancy index of multiple deprivation, per 1-quintile increment | -0.19 (-0.30, -0.08) p=0.001 |
| Maternal smoking during pregnancy | p<0.001 |
| Never | Reference |
| 1-4 cigarettes/day, n (%) | -1.07 (-1.90, -0.23) |
| 5-9 cigarettes/day, n (%) | -1.07 (-1.96, -0.18) |
| 10-19 cigarettes/day, n (%) | -0.68 (-1.38, 0.02) |
| 20+ cigarettes/day, n (%) | -1.71 (-2.92, -0.50) |
| “Healthy” diet principal component during pregnancy, per 1-SD decrement | -0.43 (-0.59, -0.27) p<0.001 |
| Gestational age at birth, weeks, no. (%) | p<0.001 |
| 38+ | Reference |
| 32-<38 | -0.07 (-0.58, 0.43) |
| <32 | -5.37 (-7.23, -3.50) |
| Birth length, cm, per 1-SD decrement | -0.73 (-0.82, -0.65) p<0.001 |
| Birth weight, kg, per 1-SD decrement | -2.41 (-2.68, -2.13) p<0.001 |
| Breastfeeding duration | p<0.001 |
| 6+ months | Reference |
| 3-<6 months | -0.07 (-0.54, 0.40) |
| 1-<3 months | -0.60 (-1.06, -0.14) |
| <1 month | -0.56 (-1.04, -0.09) |
| Never | -1.07 (-1.53, -0.62) |
| Childhood (ages 0-12 years) index of multiple deprivation, per 1-quintile increment | -0.23 (-0.36, -0.11) p<0.001 |
| Childhood (ages 6-34 months) household tobacco smoke exposure duration | p<0.001 |
| 0 hours/week | Reference |
| >0-5 hours/week | -0.28 (-0.67, 0.11) |
| >5-10 hours/week | -1.50 (-2.27, -0.73) |
| >10-20 hours/week | -1.30 (-2.05, -0.55) |
| 20+ hours/week | -0.95 (-1.71, -0.19) |
| “Healthy” diet principal component at 38 months, per 1-SD decrement | -0.23 (-0.39, -0.07) p=0.005 |
| Residential outdoor PM_2.5_ concentration, μg/m^3^, per 1-SD decrement | 0.04 (-0.16, 0.24) p=0.704 |

Model covariables: age at height-GaP assessment, sex and genotype-predicted height.

Abbreviations: ALSPAC = Avon Longitudinal Study of Parents and Children; SD = standard deviation; PM_2.5_ = particulate matter with diameter less the 2.5 micrometres.

*Supplementary Table 13*: Association of ancestry-specific polygenic height score-derived height-GaP with mortality in UKBiobank and MESA.

|  | **Hazard ratio per 1-SD height-GaP deficit (95%CI) p-value** | |
| --- | --- | --- |
|  | **UKBiobank** | **MESA** |
| All-cause death |  |  |
| Model 1 | 1.12 (1.10, 1.13) p<0.001 | 1.08 (1.02, 1.14) p=0.012 |
| Model 2 | 1.12 (1.10, 1.13) p<0.001 | 1.08 (1.02, 1.14) p=0.011 |
| Atherosclerotic cardiovascular disease death |  |  |
| Model 1 | 1.15 (1.13, 1.18) p<0.001 | 1.13 (1.00, 1.27) p=0.046 |
| Model 2 | 1.15 (1.13, 1.18) p<0.001 | 1.12 (1.00, 1.26) p=0.049 |
| Atherosclerotic coronary heart disease death |  |  |
| Model 1 | 1.25 (1.21, 1.29) p<0.001 | 1.27 (1.07, 1.51) p=0.005 |
| Model 2 | 1.25 (1.21, 1.29) p<0.001 | 1.28 (1.08, 1.52) p=0.004 |

Model 1: age, sex, principal components of genetic ancestry.

Model 2: Model 1 variables, genotype-predicted height.

Abbreviations: MESA = Multi-Ethnic Study of Atherosclerosis; SD = standard deviation.
